# Effect of community interventions for the prevention of suicide in adolescents and young adults: a scoping review

**DOI:** 10.1101/2024.10.19.24315812

**Authors:** Fernando Cabrera-Eraso, Manuela Torres-Solano, Mariana Vásquez-Ponce, María Alejandra López-Orozco, Carlos Tirado, María Kamila Arévalo, Laura Medina, Catalina Rodríguez, Valentina Ramírez Castellanos, Lina María González B.

## Abstract

**Introduction:** Suicide is a significant public health issue, especially in middle- and low-income countries, with higher rates among young people. Suicidal behavior includes suicide, suicidal ideation, attempts, and gestures, each with distinct definitions. Community-based interventions are crucial for suicide prevention, but there is limited research on their effectiveness, especially in low-resource settings. This review aims to assess and categorize such interventions for adolescents and young adults globally.

**Objective:** To assess the effect of community interventions for the prevention of suicide in adolescents and young adults.

**Methods:** A scoping review was conducted following the methodological framework of the Joanna Briggs Institute and the PRISMA-ScR extension. The goal was to examine the effectiveness of community-based interventions for suicide prevention among adolescents and young adults (15-30 years). After searching various databases, articles were independently reviewed, and selected studies were analyzed in collaborative meetings to reach conclusions regarding the validity and outcomes of the interventions.

**Results:** 2162 references were found, but only 12 studies met the eligibility criteria for the analysis phase. The studies explored various community-based suicide prevention interventions, including educational programs for parents, dialectical behavioral therapy, and culturally adapted initiatives for Indigenous populations. Key findings demonstrated reductions in suicidal thoughts and behaviors, though challenges such as small sample sizes, cultural nuances, and long-term effectiveness were noted.

## INTRODUCTION

Suicide represents a major public health problem, with a greater burden in middle- and low-income countries (1). It is understood as a self-destructive act that seeks death through a specific method (2), which could be related to suicide ideation that refers to thoughts, plans, desires or motivations to consummate; suicidal gesture defined as any self-inflicted behavior with the potential to cause harm, but with implicit or explicit evidence that the person has no intention of committing suicide (3); or to a suicide attempt which is understood as the set of behaviors initiated by the subject with the goal of dying, regardless of whether there is a medical injury (2). According to the World Health Organization (WHO), it is a situation that not only affects the person that attempts or commits it, but also the community around. Unfortunately, it is a topic often neglected because of all the myths, taboos, and stigmas around it (4).

The incidence of suicide varies according to region, age group and sex (5), with a rate higher in men than women, while self-harm is more frequent in women. The WHO estimates that every year 726 000 persons commit suicide and even more attempt it. In 2021, 73% of the cases occurred in middle- and low-income countries, and it was the third cause of death among people between 15 to 29 years old globally (6,7) . Its prevention can be addressed at an individual, community, and national level, and it is essential to adapt evidence-based strategies according to the specific needs of the population and the context, along with the transmission of clear messages and concrete actions (8).

Given the difficulty of access to health services in middle- and low-income countries, community interventions play an important role in suicide prevention. However, there are few studies that record the reality of avoiding deaths attributable to this measure from a community approach. This review aims to identify and categorize community-based interventions aimed at preventing suicide in adolescents and young adults worldwide, including their key components, strategies, and implementation settings, and to assess the effectiveness of these community-based interventions in reducing suicidal behaviors, improving mental health outcomes, and addressing risk factors among adolescents and young adults.

### OBJECTIVE

The main objective of this scoping review is to examine the community-based interventions implemented globally to prevent suicide in adolescents and young adults, and to assess the scope and diversity of the scientific evidence regarding their effectiveness, approaches, target populations, and application contexts.

## METHODOLOGY

### 1. Search strategy

A scoping review was conducted following the methodological framework described by the Joanna Briggs Institute and the Preferred Reporting Items for Systematic Reviews and Meta-Analyses extension for scoping reviews (PRISMA-ScR) (9).

As it has been mentioned, suicide is a major public health and the age groups with the highest suicide rates in 2019 were adolescents and young adults from 15 to 29 years of age (3), so it is important to demonstrate the preventive measures that are being applied to handle these statistics; as for community interventions, there are few studies that record the reality of the avoidance of deaths attributable to this measure. That been said, the guiding research question for this review was: “What is the scope, extent, and nature of evidence on the effectiveness of community-based interventions for suicide prevention among adolescents and young adults?”.

Relevant MeSH, EMTREE, and free-text terms were selected based on this research question. The searches were conducted in the following databases: MEDLINE (PubMed), EMBASE (EMBASE), Scielo, Cochrane Central Register of Controlled Trials (Ovid), LiLACS (BVS), and PsycArticles (PsycNet). Gray literature was reviewed using ProQuest Dissertations and Theses Global (ProQuest), and government reports and articles on community-based suicide prevention strategies were additionally sourced via a Google Scholar search.

The presentation of the data found, reviewed, and analyzed is presented in a Preferred Reporting Items for Systematic Reviews and Meta-Analysis extension for scoping review (PRISMA-ScR) flow diagram (Figure 1).

**Figure 1.**
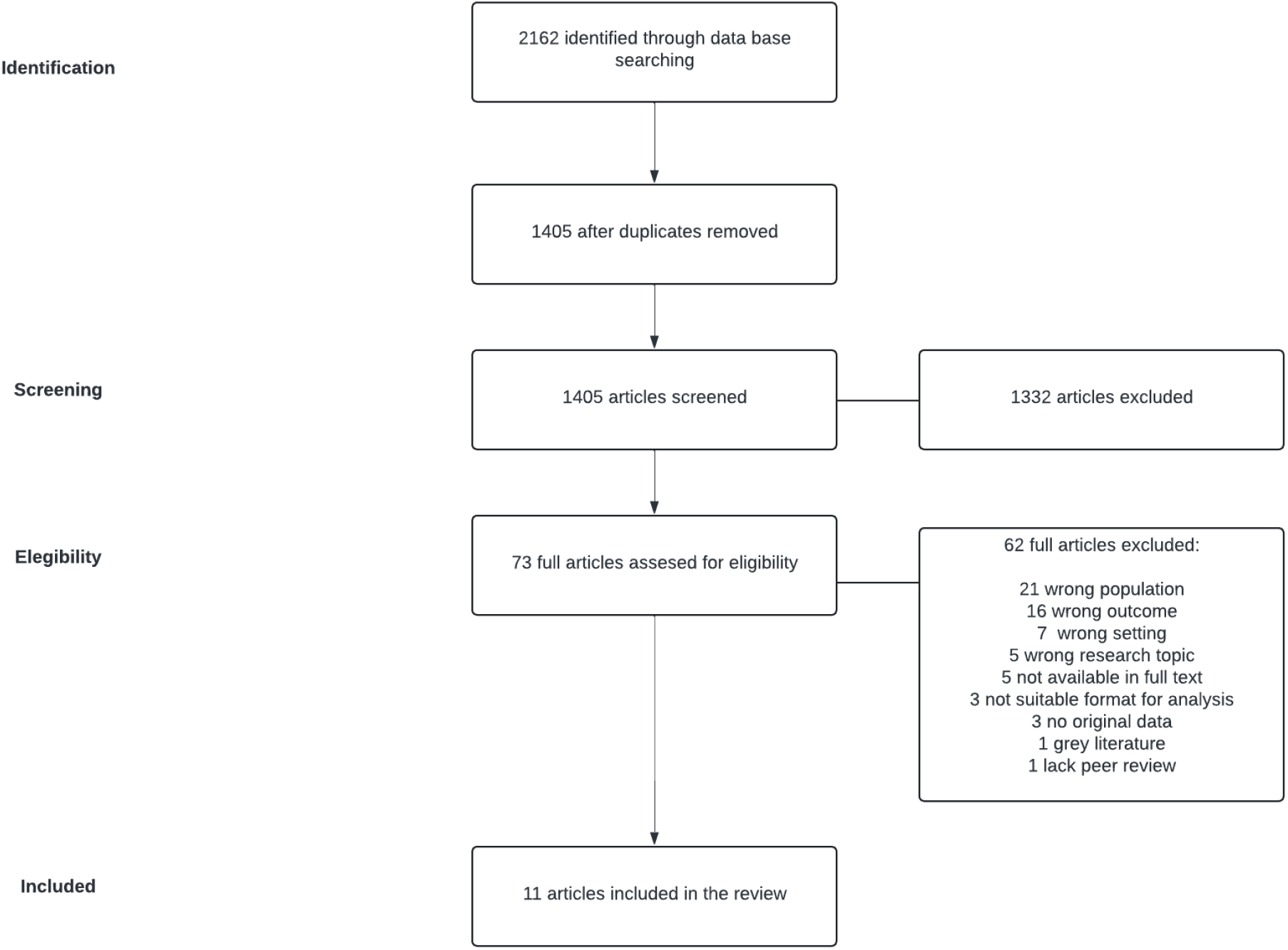
PRISMA diagram.

### 2. Eligibility Criteria

Once the literature search was completed, all 2162 retrieved citations were uploaded into Rayyan QCRI, a tool used to automatically remove duplicates. Subsequently, an initial screening was conducted based on the review of titles and abstracts by two independent reviewers. In case of disagreement, a third reviewer was consulted to determine inclusion or exclusion. Articles approved by at least two reviewers were included in the review. This finally allowed us to remove 1332 articles.

The inclusion criteria encompassed original research studies in English or Spanish, on community-based interventions that had as a reported outcome suicide prevention, including suicide, suicide attempt and suicide thoughts among adolescents and young adults between 15 and 30 years of age, including clinical trials, observational studies, and other primary, secondary, and tertiary sources, such as government reports.

### 3. Charting, Collection, Summarizing, and Analyzing data

After the initial screening phase, the full texts of the articles were read and summarized in a database with tables that included the following headings: title, article objective, population, intervention, context, type of evidence, country, participants (in the case of clinical trials), and result details. This table was used for a second review in which the researchers confirmed whether the articles met the inclusion criteria (see Appendix 1).

A new database was created with the articles that met the inclusion criteria. In addition to the previously described data, details of the interventions carried out, the evaluation tools used, the validity of the intervention, and a description of the results were included, along with the key points highlighted by each researcher for each article.

For the analysis, discussion, and conclusions, all researchers read the selected articles. The research team included one MD psychiatrist and epidemiological clinical MSc, two psychiatry residents, and seven medical students, ensuring diverse perspectives. Two virtual meetings were held, both of which were recorded and transcribed. The first phase of the review involved a rigorous assessment of the articles’ eligibility for inclusion, considering the context of the interventions, methodological approaches employed, results presented, and consensus discussions. The selected studies were further discussed in detail during a second meeting, where final conclusions were reached through a collaborative consensus process.

## RESULTS

The literature search resulted in 2162 references which were imported to Rayyan QCI. After a manual revision of duplicates, there were 1150 articles left for preliminary screening through a rigorous and blinded process in which the research team discarded articles that did not meet the selection criteria. 73 articles passed the initial screening and were assessed thoroughly for eligibility, with 12 studies that met the eligibility criteria and were employed for the analysis phase (see Appendix 2).

Two studies had a naturalistic design. Sullivant et al. (2022) (10) described an educational program directed to parents of teens, on safety considerations regarding the storage of guns and prescription drugs that could be used by adolescents to commit suicide. This intervention was developed in different community settings, including corporations and churches. A significant proportion of parents reported that they would make changes in the storage of drugs in the household and half of the participants did not report barriers in the implementation of these measures. The research team offered tools such as medicine organizers and locks for the storage of guns and medicine, which helped in the implementation of these measures. The researchers measured implementation at different phases of the program using self-developed instruments, assessing for willingness and actual changes in behaviors. Some of the barriers identified by the researchers were time to apply the measures proposed and disagreement between adults in each household.

Wood et al. (2023) (11) included 39 subjects from 16 to 24 years of age who had a history of self-harm and suicidal behaviors and their parents. They implemented group therapy with a dialectical behavioral therapy for adolescents (DBT-A) approach and the Self and Strength program with a cognitive behavioral (S&S) approach; both targeted emotional regulation and reduction of self-harm. A multidisciplinary mental health team included nurses, social workers, psychologists, and psychiatrists. The research team utilized The Suicidal Ideation Attributes Scale (SIDAS), Inventory of Statements About Self-Injury (ISAS), The Disorders of Emotion Regulation Scale –Short Form (DERS-SF), EuroQol Group (EQ5D) for measuring progress throughout the program. Participants in both groups reported a reduction of suicidal thoughts, self-harm, and emotional dysregulation; effects on quality of life were not clear in the S&S group, whereas modest improvement was reported in the DBT-A group. Some limitations identified by the research group include a small sample size, the real-life feasibility of applying such interventions due to the extension of the sessions, and the measurements were only made at the final phase for subjects that received the entirety of the interventions.

There were 3 quasi-experiment studies. Allen et al. (2009) (12) presented a social engagement prevention program catered specifically to circumpolar indigenous groups, including 54 teens and their parents. They applied a number of sessions involving the Yup’ik cultural beliefs, and developed activities adapted to the local customs with the objective of reducing youth suicidal behaviors and alcohol abuse. A manual for 26 activities, 7 directly targeted for community settings, was designed, the Qunvasgik. The research group employed Community readiness assessment, Adult Community Protective Factors Behaviours Scale and The Youth Community Protective Factors Scale to measure progress after the intervention. They found that community protective factors strengthened as adults took on more protective actions and youth felt increasingly safeguarded. This change followed a dose-response pattern, where a higher number of prevention program sessions attended by both adults and youth led to a greater improvement in protective behaviors and their sense of protection. Some strengths of this intervention include that it can be targeted to a particular community, considering the nuances of its culture and values. Some of the limitations include lack of direct measurement of suicide and alcohol abuse and a small sample size.

A single quantitative evaluation pilot study using a pre/post strategy. Barnett et al. (2020) (13) 72.9% of participants completed pre-camp surveys, whereas 62.2% completed post-camp surveys. The positive affect heightened, although exact scores were not disclosed. A sense of belonging was enhanced, especially among male participants, who indicated higher self-esteem than their female counterparts. The study’s strengths encompass its culturally pertinent design and its emphasis on psychosocial outcomes directly associated with suicide prevention, mainly through the augmentation of belonging and self-esteem. Notwithstanding the strengths and favorable outcomes, the brief duration of the intervention (five days) was identified as a disadvantage, perhaps precluding adequate time for enduring improvements. Furthermore, practical difficulties in monitoring participants from 24 isolated areas resulted in a reduced follow-up response rate, complicating the evaluation of long-term effects. The study provided significant insights into the short-term benefits of cultural interventions on Alaska Native youth, emphasizing the necessity for extended interventions and improved follow-up techniques to evaluate enduring consequences.

Walsh et al. (2020) (14) conducted a systematic review and meta-analysis encompassing 28 studies with 46,979 teenage participants enrolled in post-primary schools. The studies primarily included mixed-gender and predominantly White samples. The review examined the effectiveness of multicomponent suicide prevention interventions targeting adolescents, using a combination of universal, selective, and indicated strategies aimed at preventing suicidal thoughts and behaviors while promoting overall well-being. Interventions varied in duration, ranging from ninety minutes to two years and they used several instruments to measure outcomes, including the Columbia Suicide Severity Rating Scale, the Signs of Suicide program, and other widely used behavioral and suicide-related screening tools. Results indicated that Post-primary Suicide Prevention (PSSP) interventions were effective in reducing suicidal thoughts and behaviors in 41% of controlled trials and 71% of within-subject designs. However, the effectiveness varied based on study design and the quality of the intervention, with cluster randomized trials showing the lowest effectiveness due to challenges in school implementation. Contextual factors, such as school environment and resources, also played a significant role, with peer leader referrals being more effective in metropolitan schools. There was minimal evidence on the potential adverse effects of PSSP interventions. Limitations included the underrepresentation of lower-income countries and disadvantaged schools, a lack of diversity in the samples, and limited reporting on cross-sector collaborations and relevant mental health policies.

Among the studies analyzed in the review, Trout (2018) (15) conducted a pilot study implementing the PC CARES (Promoting Community Conversations about Research to End Suicide) intervention, a community health education and mobilization model, in Indigenous communities in native villages in Alaska. The intervention occurred in 12 villages, with populations ranging from 90 to 1,200 people, the largest being 3,200. During the intervention, local facilitators invited health workers, school staff, and community members to participate in two-hour learning circles, where information was presented through videos, case studies, and infographics about suicide prevention among Indigenous youth. The discussion format followed three key questions: “What do we know?”, “What do we think?”, and “What do we want to do?”. The intervention was measured using the PC CARES scale. The study’s results were primarily descriptive, focusing on participants’ personal experiences in the learning circles. Although active community participation was reported, no quantitative data on the effectiveness of the intervention for the target population was provided. Among the limitations is that being a qualitative and pilot study, the efficacy of the model for suicide prevention was not evaluated, nor was the specific type of population to which the intervention was applied specified.

To address mental health issues among university students, Silk (2017) (16) conducted a quasi-experimental study in the U.S. with a sample of 391 individuals aged 18 to 28 years, averaging 19.01 years old, of which 60.6% were women. The three-month intervention featured a mental health campaign with peers’ or sports celebrities’ endorsements. Following the campaign, focus groups and interviews were conducted to gather feedback on student preferences and opinions about the intervention and their likelihood of seeking counseling for peers in distress. Measurement tools assessed students’ intentions to report concerns to university counseling services, their readiness to access mental health services, attitudes toward counseling, and perceptions of mental health stigma. Results indicated increased visibility of posters in common residential areas and a statistically significant rise in students’ willingness to refer friends to counseling services. The study highlighted that nearly three-quarters of students were exposed to the UCC materials, indicating strong campaign reach. Posters and table toppers were strategically distributed at various times and locations, and emails were sent three times to reinforce the message. Although the campaign aimed to create a lasting impact with a memorable poster featuring the university’s mascot, the study faced limitations such as a quasi-experimental design without randomization, unclear research questions, and poorly defined intervention goals. The findings emphasize the need for effective campaign materials and prolonged exposure to ensure impactful results.

A separate study featured in the review was executed by Wexler (2019) (17), employing a mixed-methods technique in the U.S. that concentrated on remote Alaskan towns, comprising a sample of 112 participants and 335 non-participants. The 15-month intervention utilized the PC CARES model, designed to promote community learning about suicide prevention in collaboration with Indigenous leaders and education experts. This model included various components: community workshops and educational sessions, specific training for community leaders and facilitators, community meetings to discuss suicide prevention, and events to encourage active participation. Surveys were used before and after the interventions to measure changes in knowledge, skills, and community cohesion, and social network analysis to assess the impact on the community and the dissemination of preventive practices. Results showed significant improvements in perceived knowledge, skills, and community cohesion among PC CARES participants. PC CARES participants surpassed non-participants in knowledge and attitudes toward prevention following the intervention. Those who attended two or more learning circles showed greater improvements, indicating that increased participation enhances benefits. The social impact was also reflected in increased preventive behaviors reported by PC CARES participants, with individuals in their social networks also showing maintenance or enhancement of their preventive activities, suggesting a positive network effect. The study’s strengths included strong community engagement, cultural adaptation of the intervention, and adequate training for facilitators. Limitations included the lack of a randomized controlled trial and the absence of baseline surveys for non-participants, which restricted the comparison of results and the precise measurement of preventive behaviors.

A 2021 study by Occhipinti (18) in Australia aimed to decrease suicide rates among individuals aged 15 to 24 through multicomponent interventions, including social connectedness programs, technology-enabled coordinated care, assertive aftercare post-attempt, reductions in childhood adversity, and enhancements in youth employment. The impacts of these interventions, derived from models and simulations that account for variables such as population, psychological distress, social determinants, and suicidal behavior, were anticipated to be examined over a 20-year span. As the findings were purely projections, the results were obtained expeditiously, resulting in a brief investigative process. The findings indicated a 28.5% reduction in self-harm hospitalizations (suicide attempts) (95% confidence interval 26.3–30.8%) and a 29.3% decrease in suicide fatalities (95% confidence interval 27.1–31.5%); these reductions were primarily attributed to enhanced social connectedness and diminished social isolation. Due to the interventions being based on a large population throughout an extensive area, it was unable to tailor the program to the cultural framework and community development. At the conclusion of the program, the primary findings indicated that the most significant effects on suicidal behavior among youth are probably attained through a combination of targeted mental health and suicide prevention strategies. The constraints were the reliance on data quality and the accuracy of assumptions derived from a model.

### School interventions

An investigation conducted by Adrian in 2020 (19) using a mixed-methods approach to evaluate the deployment of Technology-enabled services (TESs) for several functions, including suicide risk management inside educational environments. In this study, school personnel analyzed the responses of students in grades 9 to 12 through an open survey and began monitoring students at risk of suicide. Utilizing a Consolidated Framework for Implementation Research, the personnel identified various determinants associated with favorable implementation outcomes of the TES. This instrument was regarded as moderately effective for suicide prevention, dependable based on the collected data, and could assist in identifying unrecognized challenges among adolescents. The primary aspects requiring attention included alignment with cultural values and norms within the educational environment, enhanced focus on confidentiality and privacy, and adaptability in student support methods.

A 1995 study by Laframboise focused on creating a suicide prevention program within the Zuni culture. The program was especially designed for the Zuni culture to determine whether the suicide prevention skills established in accordance with community specifics were more effective than general suicide prevention programs used within the community. The study evaluated a culturally compatible, school-based life skills curriculum using a multiple skills survey; however, no significant results were observed at the study’s conclusion. Despite this, the post-test evaluation conducted in 2010 indicated a positive impact on suicide prevention (20). A notable strength of the program was its foundation in and direct involvement with the culture for which it was developed. The shortcomings included the exclusion of parents from the curriculum, resulting in a misconception among them that the program was condemning their parenting and attributing higher culpability for suicide to them. Another limitation was the political aspect of the community; following a leadership change, the program was terminated because the new administration believed it had fulfilled its purpose by reducing suicide rates. The ignorance of those who had not engaged with the program from the outset and were unaware of its long-term significance undermined the program’s potential and its impact on the community.

## DISCUSSION

This review provides a comprehensive overview of the current approaches to combat this critical public health issue. Despite the lack of access to mental health resources, suicide rates continue to increase among young people in low- and middle-income countries. In this review, different types of community interventions were analyzed and evaluated for their scope, nature, and effectiveness. The research provided insights into key findings, interpretations, gaps in the literature, relevance to future research and practical applications in suicide prevention strategies.

The review uncovered several significant insights into the success of community-based interventions. Additionally, A variety of approaches are employed by these programs, ranging from parent-focused educational programs to culturally specific initiatives targeting Indigenous individuals. Despite their individuality, these approaches have a shared goal of decreasing suicidal ideation, self-harm, and completed suicides in young people by encouraging protective behaviors within communities and addressing risk factors such as emotional dysregulation, social isolation, access to lethal means.

An educational program for parents was the subject of one of the studies examined, which emphasized safety measures like securing firearms and medications, both of which are frequently used in suicide attempts. Based on a community-based intervention, numerous parents who participated in the program reported behavioral changes, such as improved storage of medications and weapons. The necessary modifications were essential in reducing the accessibility of lethal alternatives for young people who are at risk of committing suicide. The study found that while these changes were successful, they also encountered hindrances, such as some parents struggling to implement them due to time or disagreement among households.

Both the DBT-A program and the S&S program were conducted in a study that focused on emotional regulation of behavior and self-harm reduction. Both 16-24 year-olds and their parents were given interventions, which involved self-harm and suicidal behavior in adolescents (11). Among participants, there was a decline in both emotional dysregulation and suicidal ideation. However, the study encountered issues with the limited sample size and the challenge of implementing intensive therapy in real-life situations for several years. The DBT-A group demonstrated greater quality-of-life improvements compared to the S&S group, suggesting that specific therapeutic models may be more effective for certain populations.

Culturally adapted interventions were also a significant area of success. The Yup’ik Indigenous community was the focus of a quasi-experimental study that utilized culturally relevant sessions to engage both young people and their parents in reducing alcohol use and suicidal behavior. The integration of local customs and cultural beliefs in this intervention revealed a dose-effet relationship: the more sessions participants attended, the better their protective behaviors were and their sense of community protection they experienced (12). Indigenous communities require community-driven interventions that are culturally sensitive, as shown by this study, which may not align with traditional mental health programs.

These interventions highlight the need for suicide prevention to be individualized to reflect local and cultural values. Programs that recognize and integrate local values, like the Yup’ik initiative, are more likely to succeed because they encourage trust, engagement, and long-term commitment from participants. The lack of direct measurement of outcomes and small sample sizes often limit the effectiveness. This defect emphasizes the necessity of more rigorous evaluation methods to determine the long-term effectiveness of such programs.

Interventions that focus on community-based efforts can reduce suicidal behavior and ideation by involving important members of society, such as parents, educators, and local leaders. These measures highlight the need to address not only the individual at risk but also the broader social context in which they exist. Community interventions, which promote protective behaviors, improve emotional regulation, and reduce access to lethal means, can create a supportive environment that can also decrease the risk factors of suicide.

Educational initiatives that engage parents and offer practical assistance in acquiring guns and drugs demonstrate how community involvement can directly impact suicide prevention. Rather than being available in clinical settings, which are often difficult to obtain in low-resource areas, these programs aim to provide effective interventions at home. Similarly, DBT programs that focus on emotional dysregulation, one of the primary risk factors for suicide, demonstrate that community-based therapeutic interventions can result in measurable reductions in self-harm and suicidal thoughts, even in those with high susceptibility to such interventions.

Indigenous interventions that incorporate their cultural values reinforce the importance of community involvement. These are successful programs because they incorporate the cultural norms of their target populations while preserving traditional customs, values, and protective measures. Long-term success in suicide prevention is dependent on the community feeling a sense of ownership and responsibility.

While these results are positive, the review does highlight several shortcomings. A lack of long-term data on the sustainability of interventions is a common issue in studies with limited sample sizes. Additionally, these findings cannot be generalized. Additionally, cultural differences greatly influence the impact of community interventions. In rural communities, interventions may be highly effective in addressing Indigenous issues, but not in urban settings with diverse social dynamics and risk factors. This demonstrates the importance of adaptable, flexible models of intervention that are tailored to different populations.

This review identified several critical literature gaps. Firstly, there is a significant lack of research on low- and middle-income countries, even though areas with the highest suicide rates are often poor. Currently, most of the research has concentrated on countries with higher incomes and better access to mental health services. This leaves a gap in understanding how community-based interventions can be applied in resource-limited environments where traditional healthcare infrastructure may not have been available.

Most studies do not provide long-term follow-up, which is a significant gap. While many interventions show short-term reductions in suicidal behavior, it’s unclear if these effects are long-lasting. Despite the lack of data on whether these improvements persisted for months or years, DBT and S&S interventions were found to have led to reduced suicidal thoughts in the study. There is a pressing necessity for studies that can track participants over extended periods and evaluate the long-term impact of community interventions. In addition, research on how men and women can prevent suicide is lacking. Although there is a variation in suicide risk factors per gender, many studies reviewed did not provide detailed information on how interventions affect individuals with different levels of stress and anxiety. Suicide rates among men are higher than those among women, with the latter being more likely to self-harm. If we acknowledge the differences, it could lead to more focused interventions that cater specifically for each gender.

Despite the overlap between the Joanna Briggs Institute guidelines and the PRISMA-ScR framework, this scoping review is notable for its thorough methodology. The review incorporated various study designs and utilized multiple databases to document community interventions, including educational programs, therapeutic approaches, and culturally specific initiatives. This multidisciplinary approach allowed the researchers (including psychiatrists, mental health practitioners and medical students in particular) to provide very different viewpoints through which to interpret the results.

Nonetheless, the review had several limitations. Among the main issues was that few studies met eligibility standards. Only 12 out of the 2162 references were included in the final analysis. Because of this small sample size, the results cannot be generalized and need to be further researched in this regard. Furthermore, the diversity of studies included in such studies hindered the ability to make conclusive determinations about the overall impact of community-based interventions. However, studies varied greatly in both design and across different target groups and cultural contexts making comparisons and synthesis of the data difficult.

Furthermore, numerous studies utilized self-reported data, which can be biased in sensitive areas like mental health and suicide. Additionally, A lack of long-term data hindered the ability to assess the effects of interventions. This review has a significant impact on practitioners and policymakers working to prevent suicide. The importance of addressing local populations’ specific needs and contexts in suicide prevention programs is highlighted by the success stories of culturally tailored, community-driven interventions. Indigenous communities may gain the most from interventions that incorporate cultural practices and values, as demonstrated by the success of the Yup’ik program. Ensure that programs of cultural significance are funded and implemented with appropriate measures by policymakers. In addition mental health professionals should involve parents, schools and other community stakeholders in suicide prevention efforts. Programs that offer practical tools, such as those for obtaining firearms and medications, can directly reduce the risk of suicide. Education, work environments and community organizations play an essential role in identifying individuals at risk and providing early warning.

The deficiencies uncovered in this review suggest multiple directions for future research. Long-term studies are necessary to determine the effectiveness of community-based interventions in reducing suicide rates over time. In the future, research should be directed towards low- and middle-income countries, where effective suicide prevention strategies are especially important. Further investigation of gender differences in suicide risk and intervention effectiveness, as well as the impact of socioeconomic factors on outcomes, is required.

It is important to examine the feasibility of technologically based interventions in areas with limited access to mental health services in future research. Adolescents and young adults’ use of digital platforms, such as Facebook and Snapchat, is becoming increasingly important in their lives.

In summary, the findings of this scoping review indicate that community-based interventions can be effective in reducing suicidal behavior among adolescents and young adults. Although the results are positive, significant gaps exist in the literature, particularly regarding long-term effectiveness, population diversity, and low-resource research. These deficiencies must be addressed to create more effective, sustainable and relevant suicide prevention strategies around the world.

## Data Availability

All data produced in the present work are contained in the manuscript.
All data produced in the present study area available to the authors

## Appendix

Appendix 1. Results of literature search on the database used.

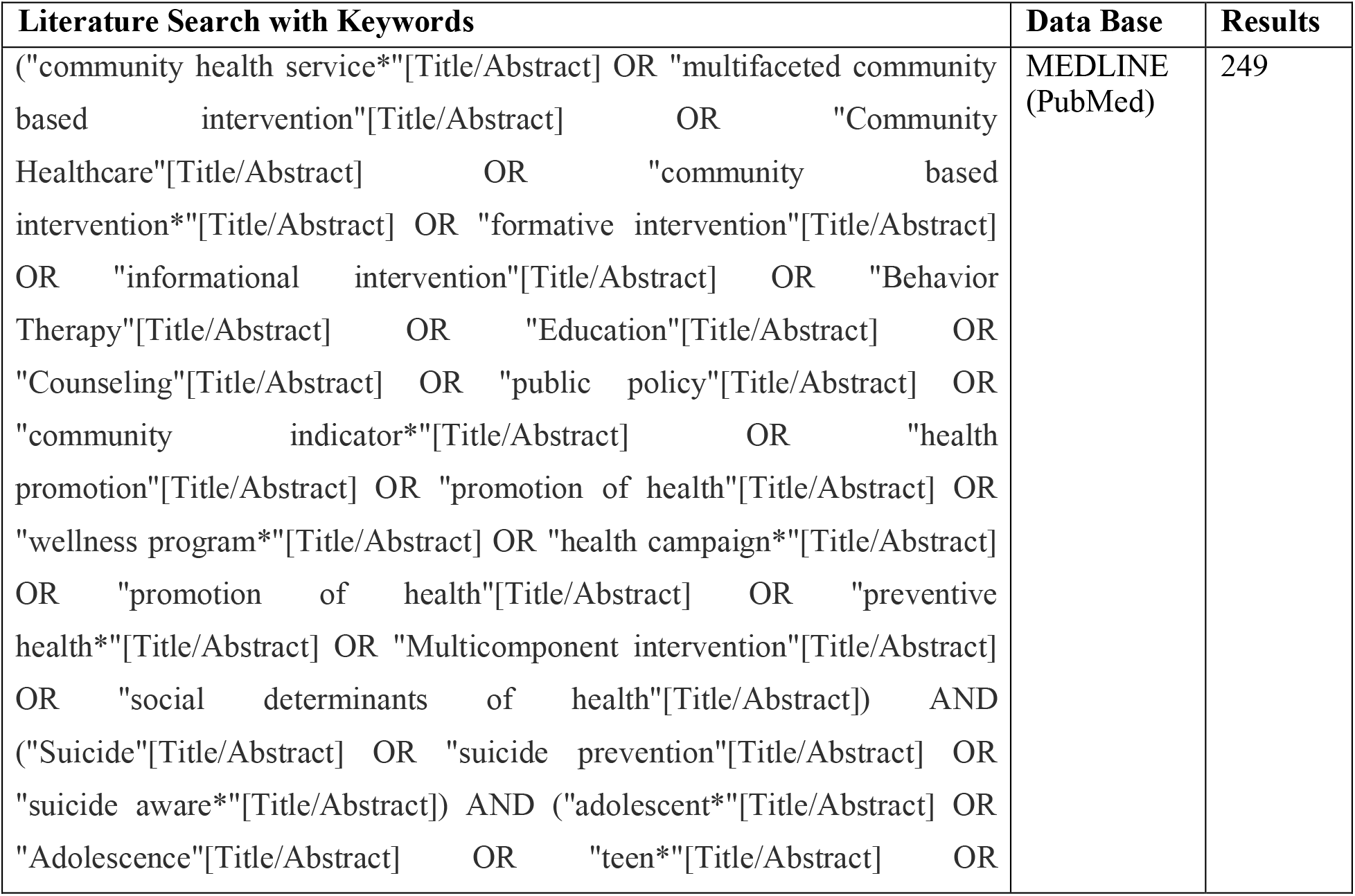
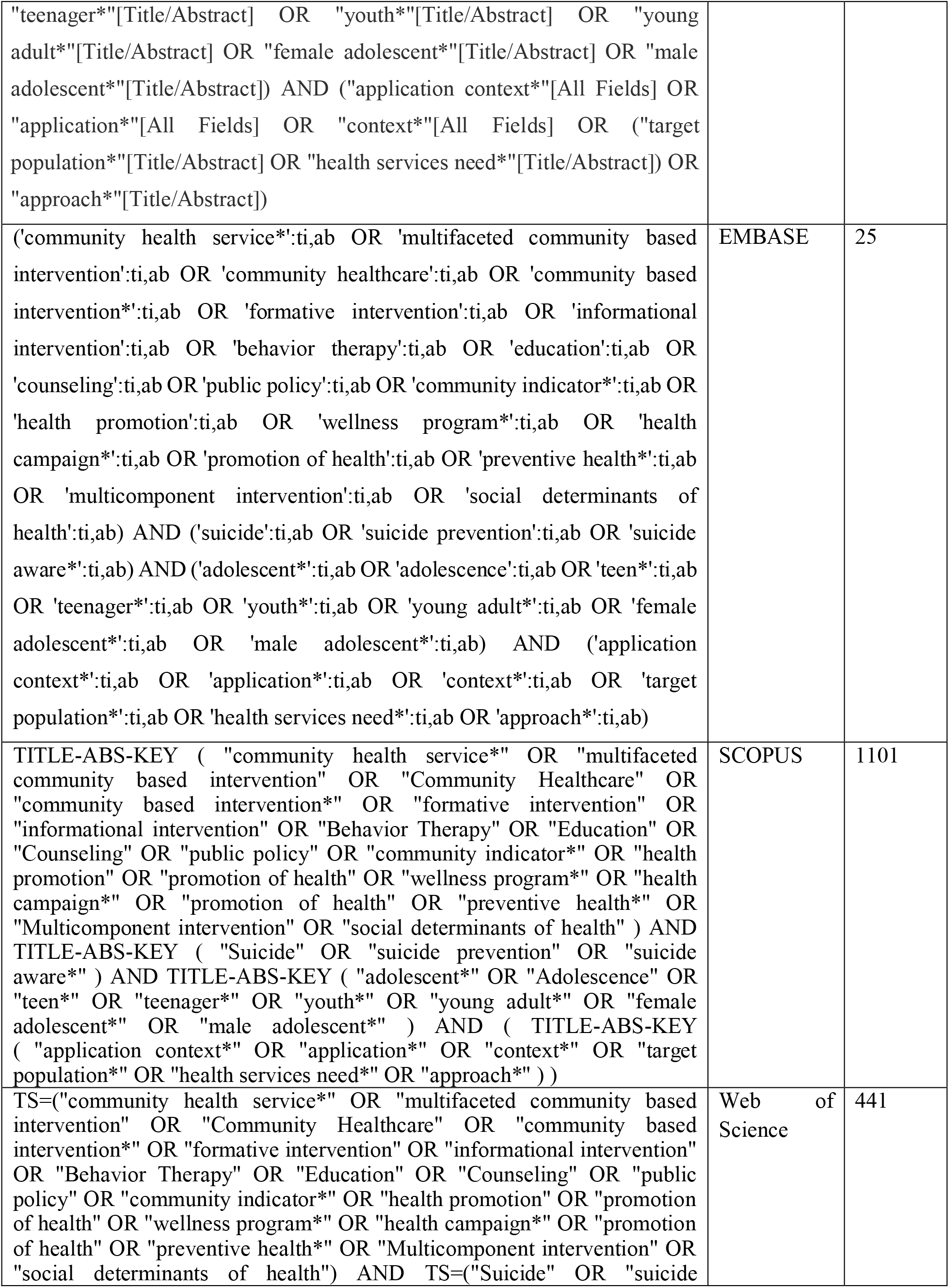
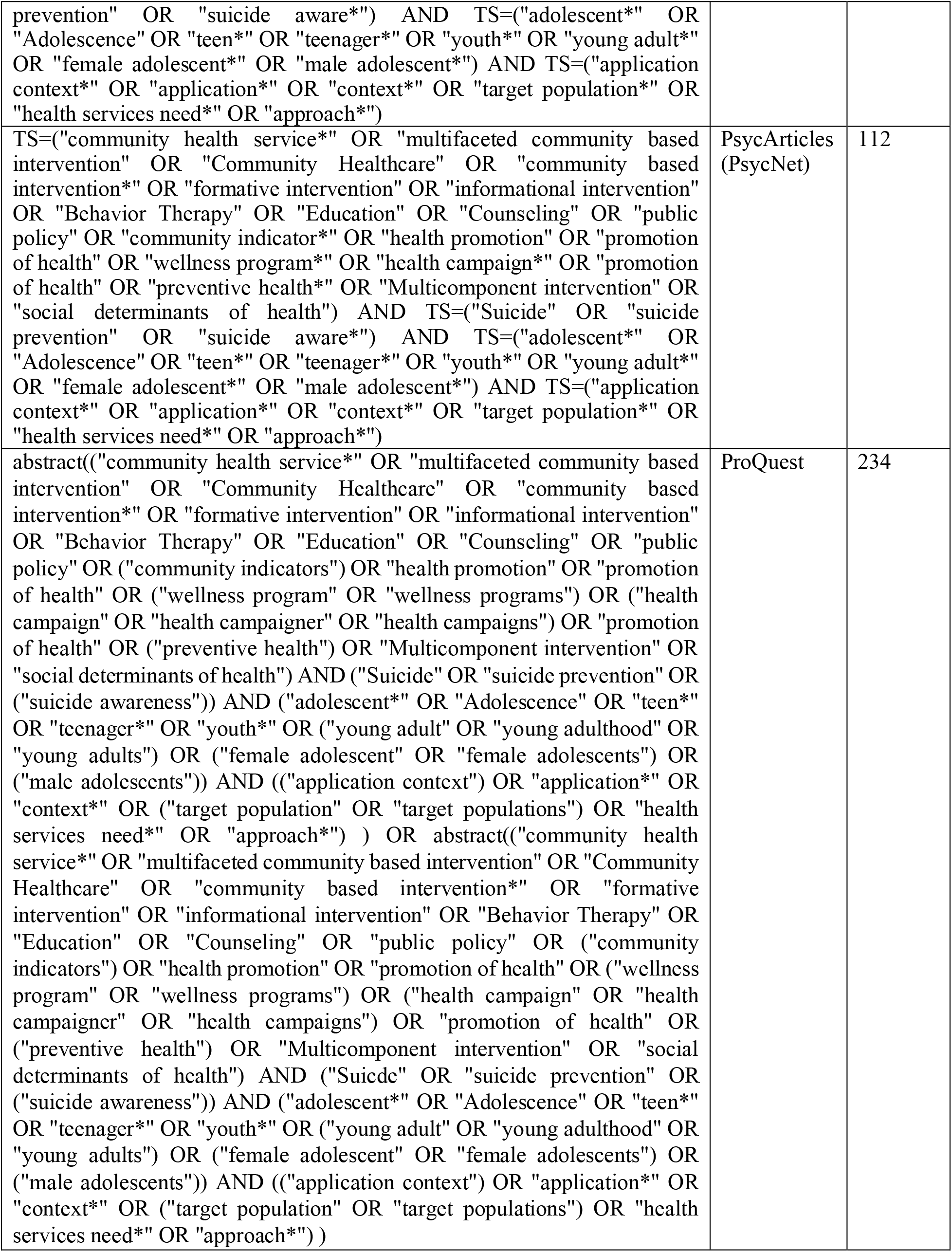

Appendix 2. Results summary.

| Title | Authors | Publication year | Design | Participants | Intervention | Key Findings |
| --- | --- | --- | --- | --- | --- | --- |
| Motivating Behavior Change in Parents for Suicide Prevention in the Midwest, USA | Sullivant, S et al | 2022 | Naturalistic study | 410 parents<br>178 < 45 years, 232 > 45 years.<br>331 women, 70 men. | Educational. Parents were presented with a presentation titled “Prepped and Ready: Parenting into the Teen Years”. The presentation was described as an opportunity for parents to learn about changes they can make to improve safety for adolescents in our community. | Most of the parents committed to embrace the safety recommendations for saving medications and guns in their households. |
| A Pilot Evaluation of Culture Camps to Increase Alaska Native | Barnett, J.D. et al | 2020 | Pilot study | 111 Alaska Native young, from 13 to 18 years. 56.75 were male, and | Educational and social engagement. They made 5-day camps, in which they expected to connect Alaska Native youth to | There was an improvement in the scores for positive affect, increased sense of belongingness |
| Youth Wellness |  |  |  | 43.25 female. | their Indigenous culture through stories, skill building, and strengthening interpersonal relationships between other youth and adult mentors and Elders. | s, and greater perceived internal ability to handle potential life stressors. |
| Decoloniality as a Framework for Indigenous Youth Suicide Prevention Pedagogy: Promoting Community Conversations About Research to End Suicide | Trout, L. et al. | 2018 | Pilot study | Participants involved people from rural and remote regions, encompassing 11 villages ranging in population from 90 to 1200, spread over approximately 40,000 square miles. | Educational. They implemented the PC CARES - Promoting Community Conversations about Research to End Suicide, which aims to improve self-determination in suicide prevention. | Self-reported from the experience of the individuals, who voiced appreciation for the process of coming together to take collective action, and for the fact that local people were leading the effort in a manner both respectful of local culture and responsive to local need. |
| Evaluation of a Social Norms Approach to a Suicide Prevention Campaign | Silk, K.J. et al. | 2017 | Quasi experiment | 391 college students, from 18 – 28 years, with a mean age of 19.01. | Educational and social engagement. They made propaganda about mental health through the campus, and then made surveys to gather information of the preferences and opinions of the college students about these types of interventions, and the possibility to reach for counseling for a friend at risk. | Students did recognize the propaganda, and they had a positive perception to reach for counselling. |
| Suicide prevention as a community development process: Understanding circumpolar youth suicide prevention through community level outcomes. | Allen, J. et al. | 2009 | Quasi experiment | 54 youth participants from Circumpolar indigenous communities (Yup'ik) 23 male and 32 female, median age 14, and 43 adults | Prevention program (Social engagement). They provided a manual (Qunvasgik) with activities that were adapted to reflect local customs and circumstances, delivered in 1 or more sessions for each activity (26 activities, 7 activities directed directly to in a community level) | Community protective factors improved as adults engaged in more protective behaviors and as youth felt more protected. This improvement followed a dose-response pattern, meaning the more prevention program sessions attended by both adults and youth, the greater the positive effect on protective behaviors and perceptions. |
| A Systematic | Walsh, E.H. et al. | 2022 | Systematic review | Adolescents 11-19 years | Multicomponent. Research on | Effectiveness Post- |
| Review of School-Based Suicide Prevention Interventions for Adolescents, and Intervention and Contextual Factors in Prevention |  |  | and meta-analysis | of age in post-primary school. 28 studies retained containing 46,979 Participants, mix gender, largely white samples or predominant from white countries | different preventive strategies: universal (targeting school populations) selective (identifying risk groups) and indicated (targeting suicide thoughts and behaviors in high risk adolescents). This intervention not only tries to identify suicidal thoughts but also includes interventions for them focusing on primary results and other well-being outcomes. | Secondary Suicide Prevention (PSSP) interventions can effectively reduce suicidal thoughts and behaviors (STBs) among adolescents, with statistically significant reductions observed in 41% of controlled trials and 71% of within-subject designs. |
| Reducing youth suicide: systems modelling and simulation to guide targeted investments across the determinants | Occhipinti, J.-A. et al. | 2021 | Pilot study. | 50 local stakeholders, including representatives from health and social policy agencies, non-government organizations, primary care providers, emergency services, research institutions, | Multicomponent. They developed a model that incorporated defined pathways from social determinants of mental health to psychological distress, mental health care, and suicidal behavior. | A combination of social connectedness programs, technology-enabled coordinated care, post-attempt assertive aftercare, reductions in childhood adversity, and increasing youth employment |
|  |  |  |  | community groups, and, importantly, people with lived experience of suicide. |  | projected the greatest impacts on SB, particularly in a youth population, reducing self-harm hospitalisations (suicide attempts) by 28.5% (95% interval 26.3–30.8%) and suicide deaths by 29.3% (95% interval 27.1–31.5%) |
| Reducing youth suicide: systems modelling and simulation to guide targeted investments across the determinants | Adrian, M. et al. | 2020 | Feasibility study. | 111 students in the 9th to 12th grade and 9 school personnel. | Multicomponent. Via an open-ended survey they evaluated implementation outcomes (acceptability, feasibility, and appropriateness) of the Technology-Enabled Service – TES in the students. The school personnel completed laboratory-based usability testing of a dashboard tracking the suicide risk of students, quantitative measures, and qualitative | implementation on determinants and outcomes suggest that both student and school personnel users view TESs for suicide prevention as moderately feasible and acceptable based on the Acceptability of Intervention Measure and Feasibility of Intervention Measure and as needing |
|  |  |  |  |  | interviews to understand key implementation outcomes and determinants | improvements in usability based on the System Usability Scale. |
| Community mobilization for rural suicide prevention: Process, learning and behavioral outcomes from Promoting Community Conversations About Research to End Suicide (PC CARES) in Northwest Alaska | Wexler, L. et al. | 2019 | Mixed methods. | On remote Alaskan towns, comprising a sample of 112 participants and 335 non-participants . | Multicomponent. The 15-month intervention utilized the PC CARES model, designed to promote community learning about suicide prevention in collaboration with Indigenous leaders and education experts. | Significant improvements in perceived knowledge, skills, and community cohesion among PC CARES participants. PC CARES participants surpassed non-participants in knowledge and attitudes toward prevention following the intervention. |
| The Zuni Life Skills Development Program: A school/community-based suicide prevention intervention | LaFromboise, T.D. et al. | 2010 | Post-test of a quasi-experiment | Adolescents of the Zuni community | Multicomponent. The program was evaluated through self-evaluation, behavioral observation and peer evaluation. | Positive impact on suicide prevention. A notable strength of the program was its foundation in and direct involvement with the culture for which it was developed. |
| Implementat<br>ion of<br>evidence-based group<br>intervention<br>s in a<br>community-based<br>mental<br>health<br>service for<br>self-<br>harming and<br>suicidal<br>youth | Wood, N.M.<br>et al. | 2023 | Naturalist<br>ic | 39 subjects<br>from 16 to<br>24 years of<br>age that had<br>history of<br>self-harm<br>and suicidal<br>behaviors<br>and their<br>parents | Multicomponent.<br>They<br>implemented<br>group therapy<br>with a dialectical<br>behavioral<br>therapy for<br>adolescents<br>(DBT-A)<br>approach and the<br>Self and Strength<br>program with a<br>cognitive<br>behavioral (S&S)<br>approach both<br>targeted on<br>emotional<br>regulation and<br>reduction of self-harm. | Participants<br>in both<br>groups<br>reported a<br>reduction of<br>suicidal<br>thoughts,<br>self-harm<br>and<br>emotional<br>dysregulation<br>, effects on<br>quality of life<br>were not<br>clear in the<br>S&S group<br>whereas<br>modest<br>improvement<br>was reported<br>in DBT-A<br>group |

